# From Registration to Insight: How STRONG AYA Transforms Registry Data to Enhance Decision-Support Tools for Adolescent and Young Adult Oncology

**DOI:** 10.64898/2026.04.03.26350064

**Authors:** N. F. (Nicola) Hughes, J. (Joshi) Hogenboom, R.N. (Robert) Carter, L.J. (Lee) Norman, V. (Varsha) Gouthamchand, O. C. (Oana) Lindner, E. (Emily) Connearn, A. (Aiara) Lobo Gomes, A. (Anna) Sikora-Koperska, M. (Magdalena) Rosińska, K. (Katarzyna) Pogoda, P. (Paweł) Wiechno, P. (Paulina) Jagodzińska-Mucha, I. (Iwona) Ługowska, S. (Simone) Hanebaum, A.L.A.J. (Andre) Dekker, W.T.A. (Winette) Van Der Graaf, O. (Olga) Husson, L.Y.L. (Leonard) Wee, R.G. (Richard) Feltbower, D. P. (Daniel) Stark

## Abstract

**Background:** Population-based cancer registers (PBCR) are important for monitoring trends in cancer epidemiology, facilitating the implementation of effective cancer services. Adolescents and Young Adult (AYA) with cancer are a patient group with a unique set of needs. The utility of PBCR in AYA is limited by the lack of AYA-specific data items. STRONG AYA, an international multidisciplinary consortium is addressing this through federated learning (FL) methodology and novel data visualisation concepts. A Core Outcome Set (COS) has been developed to measure outcomes of importance through clinical data and Patient Reported Outcomes (PROs).

We describe how data from the Yorkshire Specialist Register of Cancer in Children and Young People (YSRCCYP), a PBCR in the UK is being used within STRONG AYA and how the subsequent analyses can guide patient consultations.

**Methods:** Data from the YSRCCYP were imported into a Vantage 6 node, from which FL analyses are performed along with data provided by other consortium members. The results are extracted into the PROMPT software and integrated into patient electronic healthcare records.

**Results:** Healthcare professionals can view the results of individual PROs at various time points and in comparison, to summary analyses carried out within the STRONG AYA infrastructure. Results can be filtered by age, disease, country and stage.

**Conclusion:** We have demonstrated how a regional PBCR can contribute to a pan-European infrastructure and analyses viewed to enhance patient consultations. Such analyses have the potential to be used for research and policy-making, improving outcomes for AYA.

## Introduction

Cancer in adolescents and young adults (AYA), aged between 15-39 years, accounts for approximately 5% of new cancers diagnosed in Europe(1). Although rare, cancer remains one of the commonest causes of death in this population(2). For AYAs diagnosed with cancer, serious illness occurs at a key developmental time of life. This means that not only are they at risk of physical health implications but also delayed or non-achievement of developmental milestones including educational attainment, development of relationships and personal independence. In those that survive their cancer, the illness can result in long lasting impairment of quality of life (QoL), affecting not only on AYAs, but also on those who care for them and wider society.

Population-based cancer registers (PBCR) record all new cases of cancer within a defined population. They are important for monitoring trends in cancer incidence, mortality and survival over time, facilitating the setting of policy priorities and the planning of services. Whilst detailed research can be carried out using PBCR in common cancers, in AYA cancers the utility is often limited by the lack of AYA specific data items collected relating both to the distinct cancers common in this population, and the need to assess long-term impact on QoL.

In 2016 a report by the National Cancer Institute (NCI) AYA Cancer Progress Review Group recognised the value of pooling cohorts, including PBCR, for research purposes in AYA care(3). Further support was given by the AYA Working Group of the European Society for Medical Oncology (ESMO) and the European Society for Paediatric Oncology (SIOP Europe) who concluded that rapid solutions to ‘speak the same language’ among AYA cancer professionals was essential to improve outcomes for AYA. The STRONG AYA consortium(4) was founded in 2022 based on these principles.

STRONG AYA is a pan-European consortium comprised of AYAs with lived experience, healthcare professionals (HCPs), researchers, and policy makers who together aim to optimise data collection, management, and application for a number of purposes, including clinical decision making. Acknowledging the specific information needed to reflect the unique situation of AYA, STRONG AYA has developed a standardised Core Outcome Set (COS)(5) to measure clinical and QoL variables, in part through Patient Reported Outcomes (PROs). This COS is being collected via seven data stations across five European countries. In addition to prospective data, the STRONG AYA initiative has an inclusive perspective on re-using existing datasets: available retrospective data is harmonised to the COS, maximising data reuse(6) and enabling additional analyses. To date, this has led to the inclusion of over 26 500 AYAs from previously conducted studies, registers, and patient records. The data are stored in decentralised data stations, local to site of collection by each participating institution. Data are accessible for analysis using the modern privacy enhancing technique of Federated Learning (FL). Here only the results of analyses are exchanged, rather than centralising individual level data.

Visualisation of such European-wide analyses in clinic alongside a patient’s individual data can empower patients to take an active role in their management and provides clinicians with a valuable opportunity for personalised patient care.

In this paper, we describe how PBCR data from England, UK, were integrated within the STRONG AYA infrastructure to develop an innovative real-world hospital application. Specifically, we present an initial conceptualisation for visualising STRONG AYA analyses in conjunction with individual-level health characteristics, using the Hospital Anxiety and Depression Scale (HADS)(7) as a case study.

## Materials and methods

### Data description

The Yorkshire Specialist Register of Cancer in Children and Young People (YSRCCYP)(8) collects detailed sociodemographic and clinical information on children and young adults that have been diagnosed with cancer in the Yorkshire and Humber region since 1974. The cohort has been described previously in some detail(9) but has since extended the upper age range from 29 to 39 years to align with the international definition of AYAs. Acknowledging the importance of QoL measures in AYAs, regulatory approvals were obtained to further enhance the scope of the YSRCCYP through linkage of PRO data. Retrospective PROs flow into the YSRCCYP from the Leeds Teaching Hospital NHS Trust (LTHT) through studies led by the Patient-Centred Outcomes Research Group (PCOR) at the University of Leeds(10).

### Overcoming data inclusion challenges

The YSRCCYP’s data format is bespoke to the region and local healthcare system. In international collaborations, where harmonisation is required with other patient cohorts this can be problematic, in particular for data related to geographically specific concepts (e.g. definition of educational levels) and further compounded in FL, where individual-level data itself is not shared. In the STRONG AYA infrastructure, this challenge has been overcome by creating a shared terminology map(11) that links data items within STRONG AYA concepts to set standards. This map is used in conjunction with the Flyover tool(12,13) which maps local formats onto the STRONG AYA terminology map, producing semantically interoperable resource-description-frameworks (RDF) across the FL infrastructure. A schematic overview of this process for the YSRCCYP is provided in figure 1.

**Figure 1:**
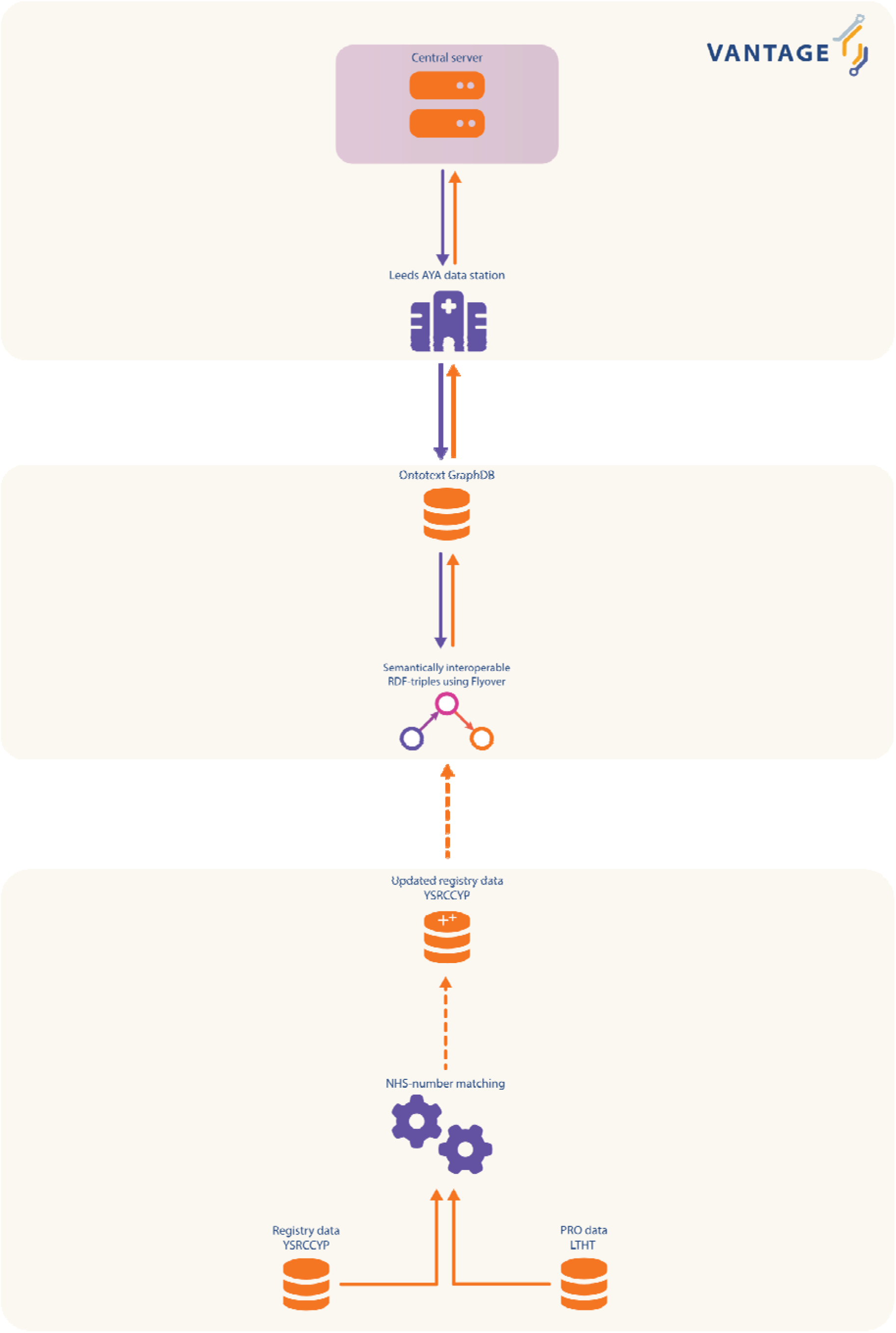
Yorkshire Specialist Register of Cancer in Children and Young People update and inclusion in the STRONG AYA federated learning ecosystem. Abbreviations: AYA: Adolescent and young adult; LTHT: Leeds Teaching Hospital NHS Trust RDF: Resource description framework; PRO: Patient reported outcome; YSRCCYP: Yorkshire Specialist Register of Cancer in Children and Young People.

### Implementation of federated analyses in a clinical setting

LTHT in collaboration with PCOR(10) co-host the software package PROMPT. Designed in-house at LTHT, PROMPT has been in use since 1999 to manage PROs and is integrated into the local electronic patient record (EPR) system. PROMPT retrieves an individual’s PROs alongside healthcare data. Traditionally PRO results are displayed with reference ranges derived from the clinical literature or instrument-defined ‘normal ranges’, but these are often historical and not AYA specific. Through STRONG AYA we sought to provide bespoke reference ranges specific to this population.

This was achieved by implementing a bridging process(14) between (i) the reference range within the database service hosted by LTHT and (ii) the STRONG AYA infrastructure, using Vantage 6(15,16). This process can periodically query the STRONG AYA data and returns values that can be used to populate the reference database.

A schematic overview of this process is illustrated in figure 2. The implementation code bridging these two services is available on <u>GitHub</u>.

**Figure 2:**
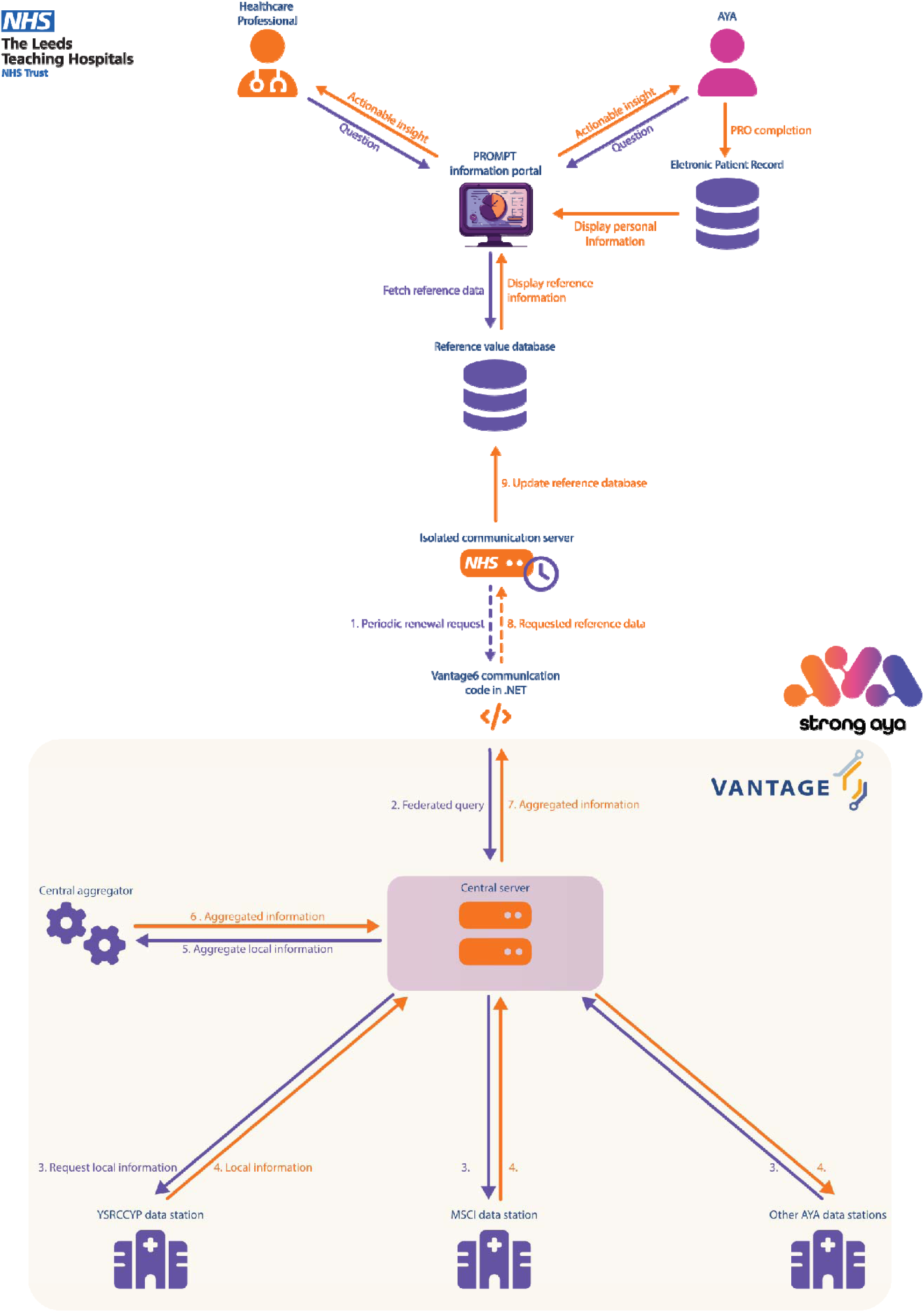
Workflow of populating the PROMPT information portal using reference data coming from the STRONG AYA ecosystem. Abbreviations: AYA: Adolescent and young adult; MSCI: Maria Skłodowska-Curie National Research Institute of Oncology; PRO: Patient reported outcome; YSRCCYP: Yorkshire Specialist Register of Cancer in Children and Young People.

For our initial use-case, we used the STRONG AYA descriptive statistics algorithm(17). This algorithm computes federated descriptive statistics including sample size counts, median and inter-quartile ranges, and allows for detailed data stratification.

The STRONG AYA descriptive statistics algorithm is open-source and available on GitHub.

Using this algorithm, we queried the STRONG AYA infrastructure for HADS data: ‘Total Anxiety Scorè and ‘Total Depression Scorè using their semantically interoperable codes – being <u>ncit:C118111</u> and <u>ncit:C118112</u> respectively(18). The configuration file used to run this query with our implementation can be found in supplementary figure 1 and are available on <u>GitHub</u>.

### Healthcare professional workshop

This work was informed by an online workshop, carried out with the purpose of determining how a portal may be used in the clinical setting both from the viewpoint of healthcare professionals (HCPs) and patients. Ten HCPs from a range of backgrounds, one scientific researcher and one AYA with lived experience attended. The workshops have previously been described(19).

## Results

### Aggregate data

The FL query revealed that 368 recordings for HADS Anxiety and Depression scales were currently available in the STRONG AYA infrastructure (table 1). Review of the STRONG AYA data management portal revealed that aggregates could be produced from two of the seven STRONG AYA partners, from existing observational research data, as shown in table 1.

**Table 1:** Retrospective data recordings included to date in the STRONG AYA infrastructure. **Asterisk** (*): Retrospective data collating is still in progress and subject to change. **Double asterisk** (**): The Netherlands Cancer Institute also contributes data of the Netherlands Cancer Registry and has national coverage. **Abbreviations**: HADS: Hospital Anxiety and Depression Scale.

| **Data station** | **Data source** | **Number of included individual AYA recordings** | **Number of HADS recordings** |
| --- | --- | --- | --- |
| *Netherlands Cancer Institute, Amsterdam (NL)* | Registry, routine-care, and research ** | 7764* | 20 |
| *The Christie, Manchester (UK)* | Routine-care | 6462 | - |
| *IRCCS National Cancer Institute Foundation, Milan (IT)* | Registry | *t.b.c\** | - |
| **<u>Yorkshire Specialist Register of Cancer in Children and Young People, Leeds (UK)</u>** | **<u>Registry and research</u>** | **<u>9095</u>** | **<u>138</u>** |
| *Centre Léon Bérard, Lyon (FR)* | Routine-care | 3045 | - |
| *Gustave Roussy, Paris (FR)* | Routine-care | 253* | - |
| *Maria Skłodowska-Curie National Research Institute of Oncology Warsaw (PL)* | Research | 210 | 210 |
| *Total* |  | 26 829* | 368 |

### PROMPT integration

As proof of concept, the HADS aggregated interquartile ranges have been visualised in PROMPT alongside a fictitious patient’s HADS scores. This is shown in figure 3 and supplementary figures 2 and 3. Here, we illustrate HADS scores over a three-month follow-up period, displayed in both a table and a graph. The table (supplementary figure 2) uses a ‘traffic light’ colour-coded system: green represents scores within the STRONG AYA reference range, and orange shows scores outside of this range. The line graph in figure 3 shows longitudinal measures for the patient alongside the reference range (represented as the green overlaid band) enabling contextualisation of the patient’s scores with those of the STRONG AYA FL analysis.

**Figure 3:**
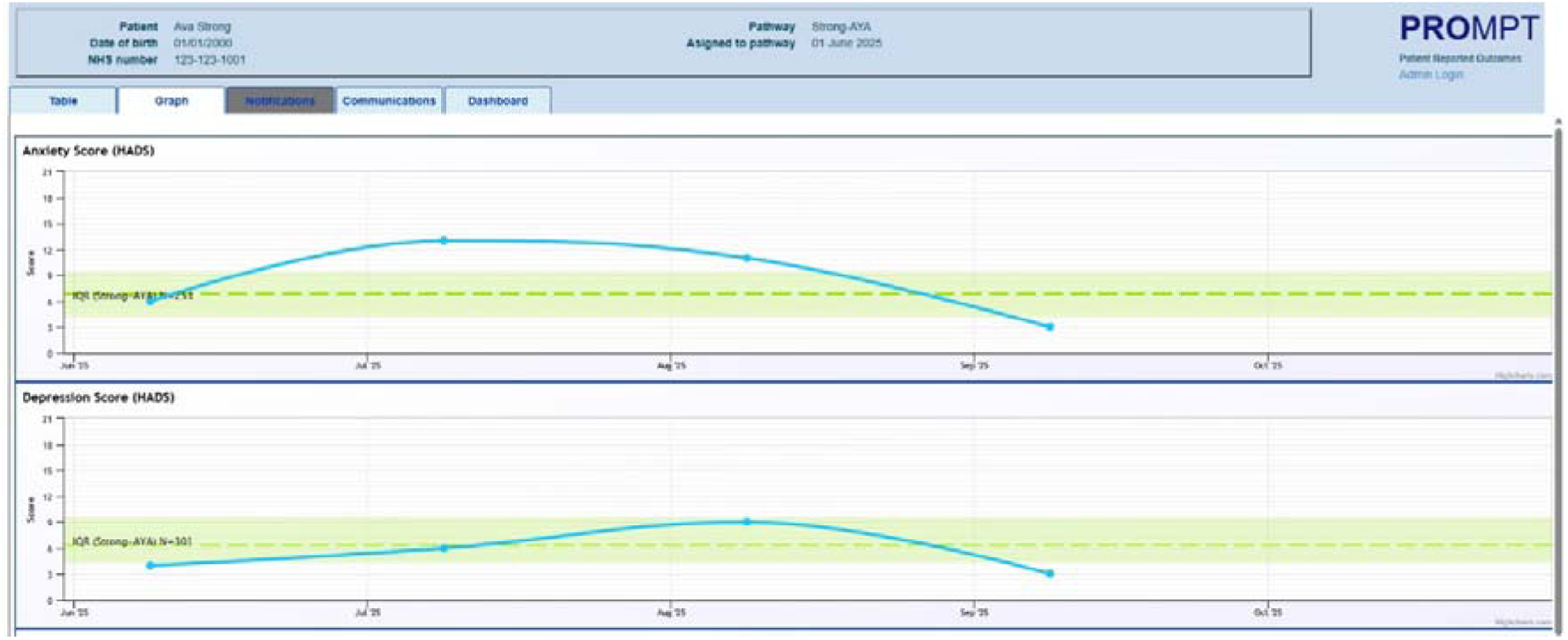
PROMPT clinician view of a patient PROMs displayed as a line graph with overlaid green band showing the aggregate interquartile Q1, median (Q2), and Q3 levels determined through federated descriptive statistics returned from the STRONG AYA infrastructure.

Extending this use of STRONG AYA-based reference values we were also able to display anxiety scores stratified by gender and for specific cancers for fictitious LTHT patients. This can be viewed as a distribution (histogram) chart using a dashboard view, mapping the patients’ value against the STRONG AYA reference ranges (supplementary figure 3).

## Discussion

We describe how regional cancer registration data can be used in FL analyses within a pan-European initiative and be visualised directly within the clinical setting to aid and direct patient care.

PROs have demonstrated clinical value in facilitating the identification and monitoring of symptoms, evaluation of long-term treatment outcomes and supporting shared decision-making(20–22). They have shown to maintain patient health and QoL and to reduce burden on healthcare services(23,24). Our intention is that cross-sectional analyses presented via the HCP portal will help AYA navigate challenges and hurdles presented to them along their treatment journey, enabling AYA to normalise their challenges against peers of the same age with the same diagnoses across Europe. Through presentation of longitudinal analyses, we aspire to enable AYAs to conceptualise how these might change over time and identify a need for intervention where appropriate.

The success and sustainability of such an application will be dependent on positive adoption by clinicians. Previously cited barriers include scepticism, unfamiliarity, time restraints and a preference for physiological measures(25). To overcome these barriers, we consulted HCPs across the consortium to ensure the product had meaningful content for users. Nonetheless, we accept that an identical portal may not be immediately applicable across all consortium members, both in terms of technological readiness and cultural barriers.

We identified that a lack of AYA-specific comparator data was a previous limitation of PROMPT. Here we provide an approach to procure dynamic and up-to-date reference range data that can be tailored to an individual’s specific case.

### Sustainability and areas for future development

Here we present a proof-of-concept for what is possible, through the STRONG AYA infrastructure and PROMPT at LTHT. The STRONG AYA infrastructure holds great potential, and across the consortium there are a number of other existing HCP and patient facing systems that can be similarly adopted to incorporate such a workflow(26–29). Whilst here we have focused on the visualisation of HADS reference scores, there are many other use cases for the infrastructure. Implementation of more complex models are possible, and our next steps include the visualisation of more sophisticated regression models including survival analyses. Cox regression modelling and Kaplan-Meier curves will enable consortium-wide survival rates to be produced enabling comparison of rates from individual centres or countries, and prognostic modelling.

Patient input and feedback is crucial to the acceptability and meaningful clinical utility of this HCP portal. Workshops are planned to obtain feedback locally at LTHT to guide ongoing development. This will ensure it is accessible and applicable to all patient groups.

The more data collected within the STRONG AYA infrastructure, the more representative, and more detailed the analyses will become. We hope that through dissemination and ongoing collaborations, the STRONG AYA initiative will be adopted internationally by other cancer centres to improve outcomes for AYAs. We acknowledge that the collection of PRO data is a time commitment for AYA. By demonstrating the potential benefits to patients of providing PROs and streamlining the collection process, we aspire to routine PROs collection becoming an integral part of the patient care pathway.

### Conclusions

Through STRONG AYA, regional PBCR data gains added value from the incorporation of a co-developed COS for AYAs and through integration with a pan-European infrastructure. This effort enables the unique data needs of AYAs to be met and facilitates research into QoL and survivorship outcomes in a safe and secure manner. By visualising such insights within clinical settings, this approach can directly inform patient care and drive shared decision-making, thus improving AYA outcomes on an international scale.

## Data Availability

The main implementation bridging from the PROMPT system to the STRONG AYA ecosystem in Vantage6 is available on GitHub: https://github.com/STRONGAYA/v6-py-client-in-dotnet, and Zenodo: 10.5281/zenodo.18483806.

https://doi.org/10.5281/zenodo.18483806

## Declaration of interest

Andre L.A.J. Dekker

- Employment: Medical Data Works B.V.
- Stock and Other Ownership Interests: Medical Data Works B.V.
- Travel, Accommodations, Expenses: Medical Data Works B.V.

Winette T.A. van der Graaf

- Research Funding: Lilly (Inst)

All remaining authors have no potential conflicts of interests to declare.

## Declaration of use of generative artificial intelligence

We used generative artificial intelligence for the following tasks: Mistral AI’s Le Chat Pro for typographical checking and Adobe Firefly for image generation (where applicable).

## Ethical approval

The Yorkshire Specialist Register of Cancer in Children and Young People (YSRCCYP) has ethical approval from the Northern and Yorkshire Multi Centre Research Ethics Committee (MREC, REC Reference: 00/3/001) and approval under section 251 of the NHS Act (2006) for holding identifiable patient data without consent from the Health Research Authority Confidentiality Advisory Group.

## Funding

J. Hogenboom, V. Gouthamchand, A. Sikora-Koperska, P. Wiechno, M. Rosińska, I. Ługowska, S. Hanebaum, A.L.A.J. Dekker, W.T.A. Van Der Graaf, O. Husson, and L.Y.L. Wee were supported in this work by the European Union’s Horizon 2020 research and innovation programme through the STRONG-AYA Initiative (Grant agreement ID: 101057482). N. F. Hughes, R. Carter, L. Norman, O. Lindner, E. Connearn, R. Feltbower, and D.P. Stark were supported in this work by Innovate UK (UK Research and Innovation) grants 10038931, 10039273, 10041045, and 1004418 under the UK government’s Horizon Europe funding guarantee. O. Husson is also supported by the Netherlands Organization for Scientific Research through a Vidi grant (ID: 198.007). L.Y.L. Wee is currently in receipt of research funds from ZonMW, Velux Stiftung and NWA-ORC.

## Prior presentation

This study and its results are original and have not been presented elsewhere and only appear as preprint on medRxiv.

## Supplementary material

**Supplementary figure 1.**
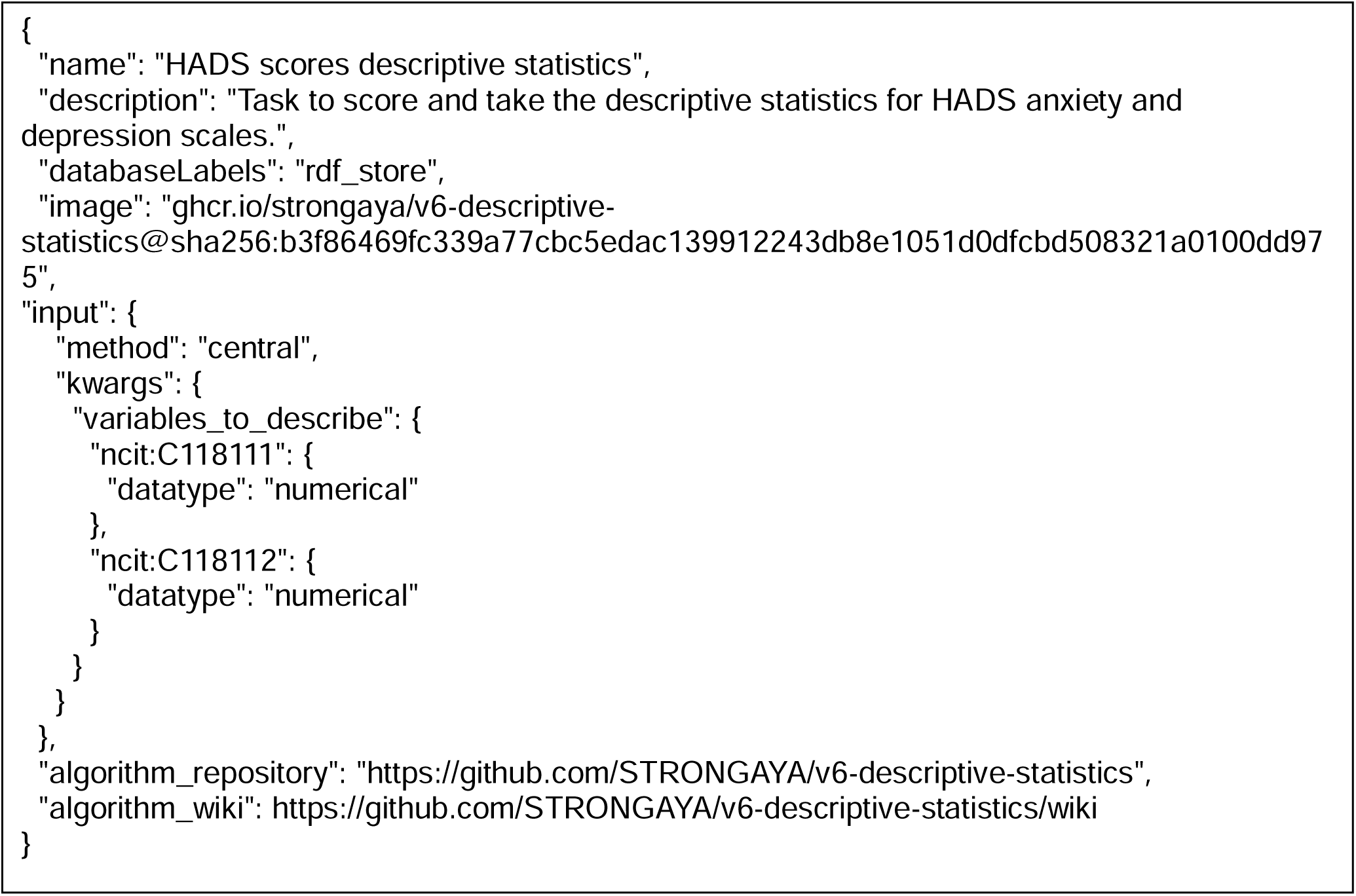
Configuration of our implementation to retrieve Hospital Anxiety and Depression Scale scores.

**Supplementary figure 2.**
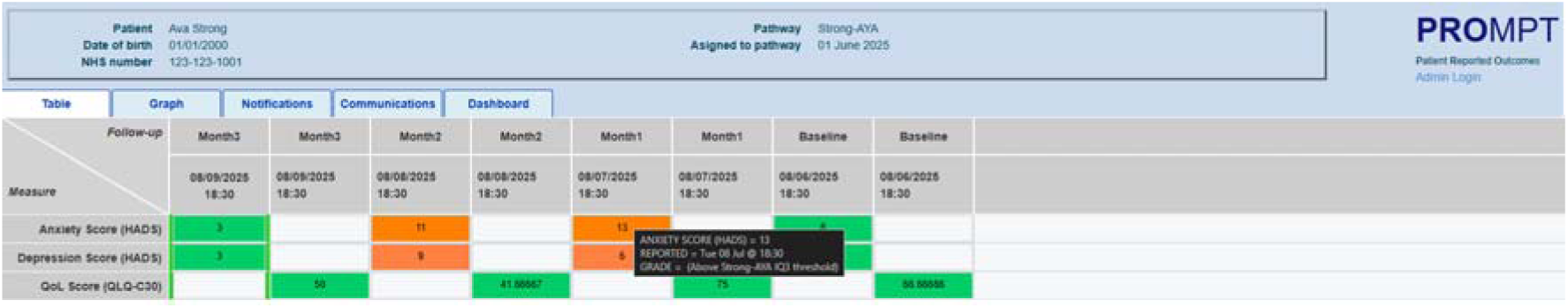
PROMPT clinician view of a patient PROMs displayed in a table with colour thresholds set to reference interquartile ranges determined through federated descriptive statistics returned from the STRONG AYA infrastructure.

**Supplementary figure 3:**
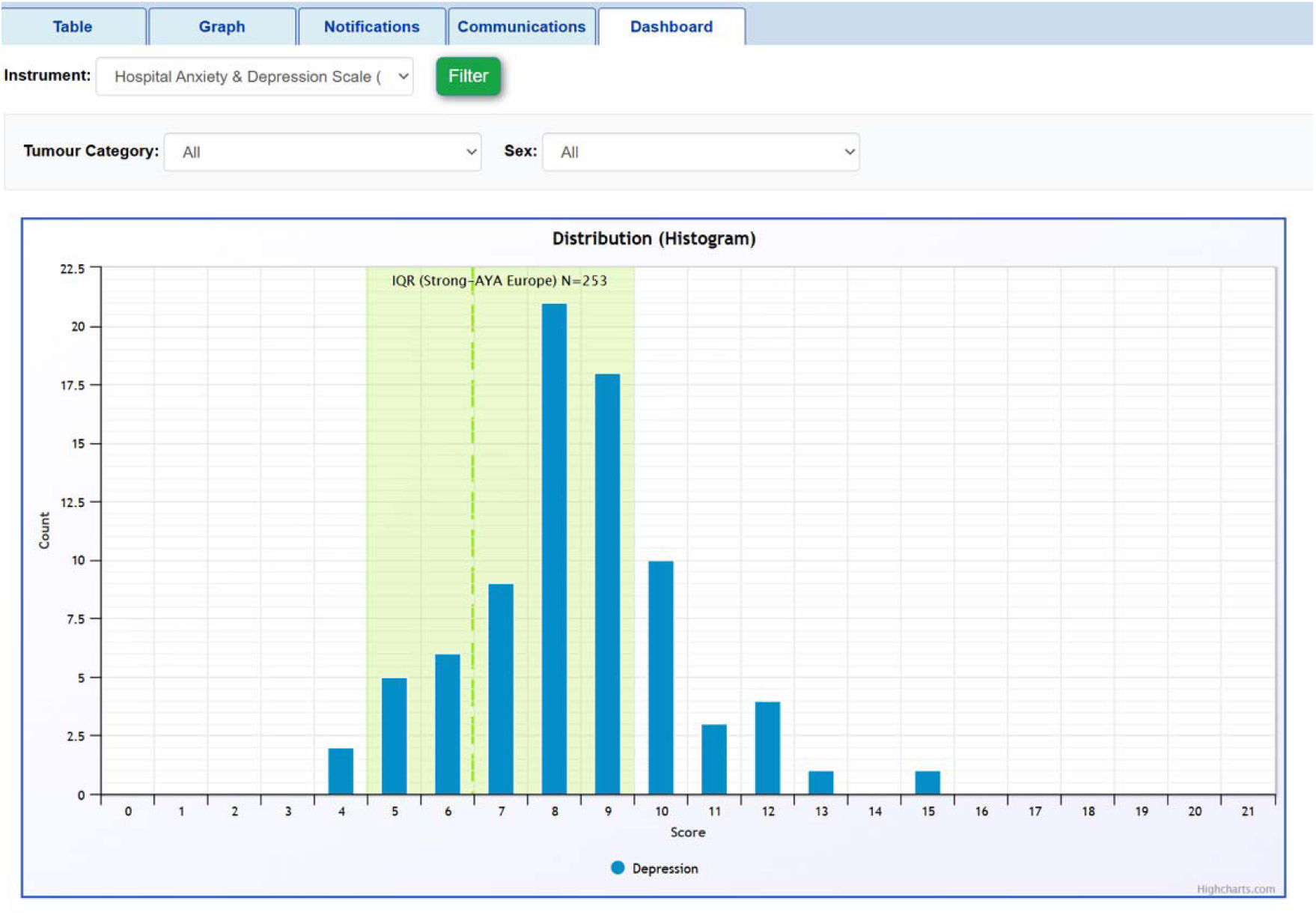
PROMPT clinician dashboard view showing a fictitious HADS score distribution for all female patients on the colorectal treatment pathway; the green background is the aggregate interquartile Q1, median (Q2), and Q3 levels determined through federated descriptive statistics returned from the STRONG AYA infrastructure.

## References

1. Trama A, Geerdes EE, Demuru E, De Angelis R, Karim-Kos HE, Troussard X, et al. Survival of European children, adolescents and young adults diagnosed with haematological malignancies in the period 2000–2013: Results from EUROCARE-6, a population-based study. Eur J Cancer. 2025 Jun;222:115336. doi:10.1016/j.ejca.2025.115336

2. Alvarez EM, Force LM, Xu R, Compton K, Lu D, Henrikson HJ, et al. The global burden of adolescent and young adult cancer in 2019: a systematic analysis for the Global Burden of Disease Study 2019. Lancet Oncol. 2022 Jan;23(1):27–52. doi:10.1016/S1470-2045(21)00581-7

3. Smith AW, Seibel NL, Lewis DR, Albritton KH, Blair DF, Blanke CD, et al. Next steps for adolescent and young adult oncology workshop: An update on progress and recommendations for the future. Cancer. 2016 Apr 5;122(7):988–99. doi:10.1002/cncr.29870

4. Horizon Europe. 2023 [Internet]. [cited 2023 Apr 26]. THE STRONG-AYA INITIATIVE: IMPROVING THE FUTURE OF YOUNG ADULTS WITH CANCER. Available from: https://cordis.europa.eu/project/id/101057482 doi:10.3030/101057482

5. Janssen SHM, van der Graaf WTA, Hurley-Wallace A, Vlooswijk C, Padilla CS, Cairns C, et al. Core Patient-Centered Outcomes for Adolescents and Young Adults with Cancer: A Comprehensive Review of the Literature from the STRONG-AYA Project. Cancers (Basel). 2025 Jan 28;17(3):454. doi:10.3390/cancers17030454

6. Sinaci AA, Núñez-Benjumea FJ, Gencturk M, Jauer ML, Deserno T, Chronaki C, et al. From Raw Data to FAIR Data: The FAIRification Workflow for Health Research. Methods Inf Med. 2020 Jun 3;59(S 01):e21–32. doi:10.1055/s-0040-1713684

7. Snaith RP, Zigmond AS. The hospital anxiety and depression scale. BMJ. 1986 Feb 1;292(6516):344.1–344. doi:10.1136/bmj.292.6516.344

8. Leeds Institute of Cardiovascular and Metabolic Medicine U of Leeds. Yorkshire Specialist Register of Cancer in Children and Young People. [Internet]. [cited 2018 Sep 20]. Available from: http://medhealth.leeds.ac.uk/info/545/yorkshire_specialist_cancer_register/

9. Cromie KJ, Crump P, Hughes NF, Milner S, Greenfield D, Jenkins A, et al. Data Resource Profile: Yorkshire Specialist Register of Cancer in Children and Young People (Yorkshire Register). Int J Epidemiol. 2023 Feb 8;52(1):e18–26. doi:10.1093/ije/dyac195

10. University of Leeds. Patient Centred Outcomes Research [Internet]. [cited 2025 Dec 17]. Available from: https://pcor.org.uk/

11. Hogenboom J, Gouthamchand V, Cairns C, Janssen SHM, Way K, Dekker ALAJ, et al. Knowledge Representation of a Multicenter Adolescent and Young Adult Cancer Infrastructure: Development of the STRONG AYA Knowledge Graph. JCO Clin Cancer Inform. 2026 Jan;(10). doi:10.1200/CCI-25-00177

12. Gouthamchand, V., Hogenboom, J., Hendriks, T., Wee, L., & van Soest, J. (2026). Flyover (v3.0.2). Zenodo. 10.5281/zenodo.18484469

13. Gouthamchand V, Choudhury A, Hoebers FJP, Wesseling FWR, Welch M, Kim S, et al. Making head and neck cancer clinical data Findable-Accessible-Interoperable-Reusable to support multi-institutional collaboration and federated learning. BJR|Artificial Intelligence. 2024 Mar 4;1(1). doi:10.1093/bjrai/ubae005

14. J. Hogenboom. STRONGAYA/v6-py-client-in-dotnet: Vantage6 client in .NET (v1.0.1). Zenodo. [Internet]. 2026 [cited 2026 Feb 23]. Available from: 10.5281/zenodo.18483807

15. Smits D, Van Beusekom B, Martin F, Veen L, Geleijnse G, Moncada-Torres A. An Improved Infrastructure for Privacy-Preserving Analysis of Patient Data. In. 2022. doi:10.3233/SHTI220682

16. Martin, F., Beusekom, B., Leurs, R., Sieswerda, M., Soest, J., Alradhi, H., Moncada-Torres, A., Baccinelli, W., Smits, D., Sanchez Gomez, L., & Harms, A. (2026). vantage6 (version/4.13.6). Zenodo. 10.5281/zenodo.18378157

17. J. Hogenboom. STRONGAYA/v6-descriptive-statistics: Vantage6 Descriptive Statistics (v2.0.0-pre) [Internet]. 2025 [cited 2026 Feb 23]. Available from: 10.5281/zenodo.17178099

18. J. Hogenboom. STRONGAYA/AYA-cancer-semantic-map: STRONG AYA semantic map (v1.0.0). Zenodo [Internet]. 2025 [cited 2026 Feb 23]. Available from: 10.5281/zenodo.17521900

19. Hogenboom J, Košir U, Deželak T, Lindner O, Hughes N, Connearn E, et al. Development of the STRONG-AYA federated learning-based adolescent and young adult cancer information portal: a co-creation process with adolescents and young adults with lived experience of cancer (Preprint). 2025. doi:10.2196/preprints.85870

20. Haverman L, Luijten MAJ, Blackford AL, Absolom K, Basch EM, van Rossum MAJ, et al. Truth and dare: patients dare to tell the truth when using PROMs in clinical practice. Quality of Life Research. 2024 Dec 3;33(12):3299–307. doi:10.1007/s11136-024-03772-3

21. Santana MJ, Haverman L, Absolom K, Takeuchi E, Feeny D, Grootenhuis M, et al. Training clinicians in how to use patient-reported outcome measures in routine clinical practice. Quality of Life Research. 2015 Jul 15;24(7):1707–18. doi:10.1007/s11136-014-0903-5

22. Gibbons C, Porter I, Gonçalves-Bradley DC, Stoilov S, Ricci-Cabello I, Tsangaris E, et al. Routine provision of feedback from patient-reported outcome measurements to healthcare providers and patients in clinical practice. Cochrane Database of Systematic Reviews. 2021 Oct 12;2021(10). doi:10.1002/14651858.CD011589.pub2

23. Takeuchi EE, Keding A, Awad N, Hofmann U, Campbell LJ, Selby PJ, et al. Impact of Patient-Reported Outcomes in Oncology: A Longitudinal Analysis of Patient-Physician Communication. Journal of Clinical Oncology. 2011 Jul 20;29(21):2910–7. doi:10.1200/JCO.2010.32.2453

24. Aiyegbusi OL, Hughes SE, Peipert JD, Schougaard LMV, Wilson R, Calvert MJ. Reducing the pressures of outpatient care: the potential role of patient-reported outcomes. J R Soc Med. 2023 Feb 9;116(2):44–64. doi:10.1177/01410768231152222

25. Fontaine G, Poitras ME, Sasseville M, Pomey MP, Ouellet J, Brahim LO, et al. Barriers and enablers to the implementation of patient-reported outcome and experience measures (PROMs/PREMs): protocol for an umbrella review. Syst Rev. 2024 Mar 26;13(1):96. doi:10.1186/s13643-024-02512-5 PubMed PMID: 38532492.

26. Kuijpers W, Groen WG, Oldenburg HS, Wouters MW, Aaronson NK, van Harten WH. eHealth for Breast Cancer Survivors: Use, Feasibility and Impact of an Interactive Portal. JMIR Cancer. 2016 May 10;2(1):e3. doi:10.2196/cancer.5456

27. Kuijpers W, Groen WG, Oldenburg HS, Wouters MW, Aaronson NK, van Harten WH. Development of MijnAVL, an Interactive Portal to Empower Breast and Lung Cancer Survivors: An Iterative, Multi-Stakeholder Approach. JMIR Res Protoc. 2015 Jan 22;4(1):e14. doi:10.2196/resprot.3796

28. Nederlands Kanker Institute. MijnAVL - Your personal patient portal [Internet]. [cited 2026 Feb 23]. Available from: https://www.avl.nl/en/mijnavl/

29. Léon Bérard Center. myCLB [Internet]. [cited 2026 Feb 23]. Available from: https://myclb.sante-ra.fr/Home/Login/tabid/229/Default.aspx?returnurl=%2f

